# Variability of Salivary and Nasal Specimens for SARS-CoV-2 Detection

**DOI:** 10.1101/2020.10.07.20208520

**Authors:** Yukari C. Manabe, Carolyn Reuland, Tong Yu, Razvan Azamfirei, Taylor Church, Diane M. Brown, Thelio T. Sewell, Justin P. Hardick, Paul W. Blair, Chris D. Heaney, Andrew Pekosz, David L. Thomas, on behalf of the Ambulatory COVID Team

**Author notes:** Corresponding Author: Yukari C. Manabe, MD, 1830 E. Monument Street, Rm 443, Baltimore, MD 21287.

## Abstract

In a large cohort of ambulatory confirmed COVID-19 patients with multiple self-collected sample time points, we compared 202 matched nasal-oropharyngeal swabs and oral salivary fluid sample pairs by RT-PCR. Nasal-oropharyngeal swabs were more sensitive than this salivary sample type (oral crevicular fluid) suggesting that not all saliva sample types have equivalent sensitivity. However, all samples that were Vero E6-TMPRSS2 cell culture positive (e.g., infectious virus) were also oral fluid RT-PCR positive suggesting that oral fluid may find the patients most likely to transmit disease to others.

## Background

There is an urgent need to improve COVID-19 diagnostics that rely on collection of nasopharyngeal specimens by health care personnel using special swabs. Salivary specimen sampling circumvents the swab supply chain bottlenecks, can be easily self-collected even by children, and is less likely to create aerosols during collection. In a hospitalized cohort of matched nasopharyngeal (NP) swab and saliva specimens, saliva specimens had a significantly higher mean log copies per milliliter of SARS-CoV-2 RNA than NP swab specimens.(1)

## Objective

We sought to determine how salivary specimens would perform if self-collected in a cohort of ambulatory COVID-19 adults.

## Methods and Findings

We compared SARS-CoV-2 RT-PCR results in 202 matched self-collected mid-turbinate nasal-oropharyngeal (OP) swabs and oral crevicular fluid collected by 67 non-hospitalized, consenting COVID-19 adults ≥18 years. (**Figure 1**) Among the samples that were concurrently positive, nasal-OP swab sample RNA measures were significantly higher (lower cycle thresholds) than oral fluid values, median 15.98 (IQR 13.96-21.21) vs 21.81 (IQR 17.35-25.27) (p-value<0.001). Of the matched samples for which at least one sample was positive, only 11.6% of samples (14/121) had higher viral burdens measured in oral fluid compared to nasal-OP swab samples. Midway through enrolment, participants were asked to add spit from the back of the mouth and throat into the Oracol collection tube to evaluate if that might enhance sensitivity. Oral fluid samples supplemented with spit remained statistically less sensitive for viral RNA detection than nasal-OP samples (p-value <0.001). Interestingly, of the 52 pre-spit samples, 4 (7.7%) had lower Ct values than nasal-OP, and, in the post-spit samples, this percentage increased to 14.5% (10 of the 69 samples). Oral fluid samples that were obtained earlier in the course of infection (≤5 days) were more likely to be positive. We cultured all RT-PCR positive nasal-OP specimens on VeroE6 TMPRSS2 cells. All culture positive samples were obtained within 11 days of symptom onset. In all matched samples in which SARS-CoV-2 was cultured, SARS-CoV-2 RNA was detected in both nasal-OP and oral fluid samples.

**Figure 1:**
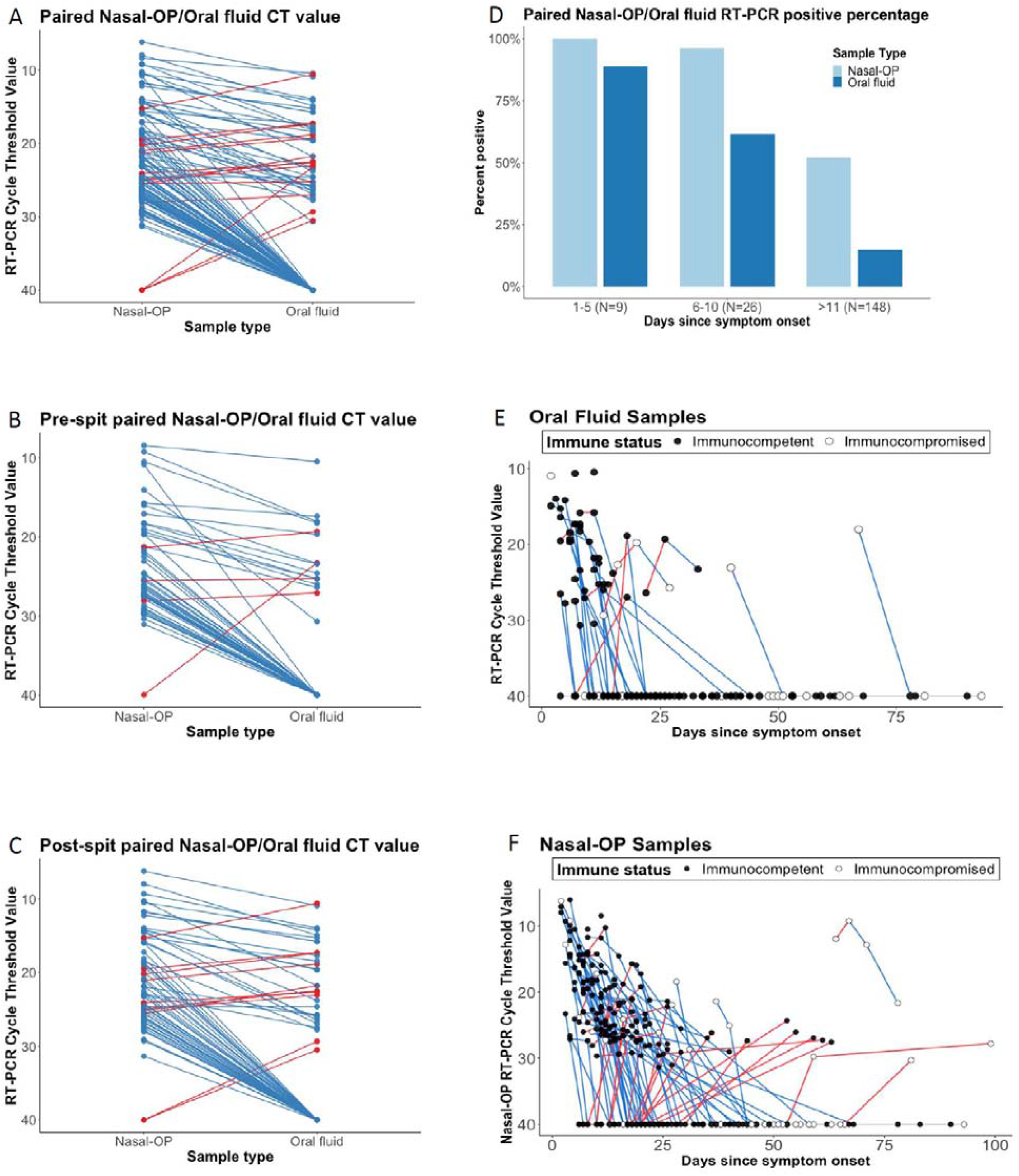
Cycle thresholds (Ct) are plotted for matched nasal-oropharyngeal (OP) swab and oral fluid RT-PCR. Viral burdens that were higher in nasal-OP or oral fluid are shown in blue and red lines, respectively in **A)** all matched specimens, **B)** oral fluid only, and **C)** oral fluid plus the addition of spit. **D)** The proportion of nasal-OP and oral fluid matched specimens that were positive in participants who are 1-5 days, 6-10, and more than 11 days after symptom onset in samples from participants where the date of symptom onset could be determined. **E)** Ct values are shown for individual participants over time. Blue lines denote decreasing viral burden whereas red lines represent increasing viral burden with increasing number of days after symptom onset in oral fluid, and **F)** nasal-OP specimens. RT-PCR Ct values for immunocompetent participants are shown in black filled circles, and for immunocompromised participants in open circles. For figures 1A-C, matched samples that were negative in both sample types were omitted.

## Discussion

Our data underscore the limitations of salivary testing for SARS-CoV-2 and challenge of COVID-19 diagnostics. In contrast to the study of Wyllie et al., we generally detected less SARS-CoV-2 RNA in oral fluid compared to nasal-OP specimens in our outpatient cohort. Others also have reported testing discordance including lower sensitivity in saliva compared to NP samples.(2) Differences in salivary collection processes might explain the findings. Wyllie et al. collected expectorated samples in the morning, a process that might increase viral abundance by enrichment of deeper samples, especially in hospitalized patients with pneumonia.(3) In the present study, samples were self-collected in an ambulatory setting using a device that was optimized for detection of oral crevicular fluid antibodies.

Oral fluid, useful for the detection of antibodies, may dilute the salivary sample and decrease its sensitivity for viral RNA detection. Many *in vitro* devices that are currently being tested for the direct detection of SARS-CoV-2 are proposing saliva including passive drool, spit, oral fluid, and sputum from clearing the throat. Not all salivary samples may be equivalent in terms of diagnostic utility and more data are needed to inform device manufacturers. Differences in the stage of infection may also factor since limited data suggest salivary tests have higher sensitivity in the first week of symptoms.(1, 4) The timing of collection, time after onset of symptoms, hospitalized vs outpatient populations, and the volume of saliva/spit may all be important for optimizing sensitivity of the saliva sample. *In vitro* device manufacturers will need to consider these factors when considering what sample types to test and when assessing assay performance.

However, our data and others consistently demonstrate the potential pragmatic use of salivary testing.(5) There was a high rate of acceptability for self-collection of oral fluid in an ambulatory setting. In addition, we and others have found that salivary sampling was 100% successful in detecting SARS-CoV-2 RNA from persons with detectable infectious virus, the widely accepted correlate of transmissibility. Thus, even if there are small reductions in analytical sensitivity, for the public health objective of identification of persons who might transmit SARS-CoV-2, home collection of salivary/oral fluid samples is an important advance that might be added to other measures like rapid testing of antigen (vs RNA) to enhance the effectiveness and scale of COVID-19 diagnostics.

## Data Availability

De-identified data is not currently available, but can be made available upon request.

## References

1. Wyllie AL, Fournier J, Casanovas-Massana A, Campbell M, Tokuyama M, Vijayakumar P, et al. Saliva or Nasopharyngeal Swab Specimens for Detection of SARS-CoV-2. N Engl J Med. 2020.

2. Sapkota D, Søland TM, Galtung HK, Sand LP, Giannecchini S, To KKW, et al. COVID-19 salivary signature: diagnostic and research opportunities. J Clin Pathol. 2020.

3. Hung DL, Li X, Chiu KH, Yip CC, To KK, Chan JF, et al. Early-Morning vs Spot Posterior Oropharyngeal Saliva for Diagnosis of SARS-CoV-2 Infection: Implication of Timing of Specimen Collection for Community-Wide Screening. Open Forum Infect Dis. 2020;7(6):ofaa210.

4. To KK, Tsang OT, Yip CC, Chan KH, Wu TC, Chan JM, et al. Consistent Detection of 2019 Novel Coronavirus in Saliva. Clin Infect Dis. 2020;71(15):841–3.

5. Azzi L, Carcano G, Gianfagna F, Grossi P, Gasperina DD, Genoni A, et al. Saliva is a reliable tool to detect SARS-CoV-2. J Infect. 2020;81(1):e45–e50.

