## Supplemental methods for "Variability of Salivary and Nasal Specimens for SARS-CoV-2 Detection"

**Supplemental Material:**

**Methods**

*Study Cohort*

Non-hospitalized adults who had received testing for SARS-CoV-2 RT-PCR from the Johns Hopkins Medical Microbiology laboratory and who were notified of their positive test were approached for participation using a verbal consent script. Inclusion criteria were age ≥18 years, able to receive study materials without violating quarantine, able and willing to perform self-collection of specimens. Participants who were able to give an oral informed consent after documentation of understanding were enrolled in the study.(1)

Immunocompromised participants included those with prior solid organ or allogeneic stem cell transplantation, current hematologic malignancy, neutropenia (ANC < 500x10^6^ cells/µL), chronic use of immunosuppressant medications, moderate to high doses of prednisone (>20 mg), recent chemotherapy, recent treatment with immune active monoclonal antibodies, and/or HIV infection with CD4 T cells<200 cells/µL.

This study was approved by the Institutional Review Board of the Johns Hopkins University School of Medicine.

*Specimen Collection*

For viral load monitoring, self-collected mid-turbinate nasal and oropharyngeal swabs were obtained after participants were given both written as well as verbal instructions. Both swabs were placed in the same 3ml viral transport medium (VTM), snapped off and capped. All samples were immediately placed in a labeled biohazard transport bag with absorbent gauze, placed upright in a clear plastic sample jar that could be washed with soapy water before being placed in the participant’s freezer. Study coordinators observed participants by video call when possible and recorded their assessment of the quality of self-collection. Mid-turbinate and oropharyngeal swabs, and Oracol (Malvern Medical Developments Ltd., Worcestershire, UK) specimens were obtained on the day that the participant received the sample box (day 0), and then 3, 7, 14 after which all samples were shipped back with gel packs. There was a final in-person collection between day 28-60 when a clinician-collected NP swab was tested on the same molecular platform. Clinical information was collected using a standardized Flu-PRO(2) instrument on the same days as sample collection, in addition to patient history in a predesigned database.

*Specimen Testing*

The VTM was aliquoted into multi-Collect tubes (Abbott Molecular, Des Plaines, Il) in 600 µl volumes. Nucleic acid was extracted from the multi-Collect tubes utilizing the Abbott Molecular m2000sp, followed by amplification and analysis on the Abbott m2000rt, both extraction and amplification were performed per manufacturer instructions. A positive reaction was defined as a reaction having a CN<31.5. When quantities were sufficient, 200 µl volumes of oral fluid were also tested.

VeroE6-TMPRSS2 (3) cell culture model was used to assess viable virus when incubated with VTM. SARS-CoV-2 specific growth was verified by indirect immunofluorescence for SARS-CoV-2 antigen (nucleocapsid and spike proteins).

*Statistical analysis* Analyses were performed using R 3.6.2 statistical software. Median CT value and corresponding interquartile range (IQR) for concordantly positive pairs were calculated for both nasal-OP and oral fluid samples. Difference of CT value between nasal-OP and oral fluid samples in matched pairs were tested using Wilcoxon signed-rank test in all samples, pre-spit samples and post-spit samples. The 0.05 significance level was used.

**References:**

1.	Blair PW, Brown DM, Jang M, Antar AA, Keruly JC, Bachu V, et al. The clinical course of COVID-19 in the outpatient setting: a prospective cohort study. medRxiv. 2020:2020.09.01.20184937.

2.	Powers JH, Guerrero ML, Leidy NK, Fairchok MP, Rosenberg A, Hernández A, et al. Development of the Flu-PRO: a patient-reported outcome (PRO) instrument to evaluate symptoms of influenza. BMC Infect Dis. 2016;16:1.

3.	Matsuyama S, Nao N, Shirato K, Kawase M, Saito S, Takayama I, et al. Enhanced isolation of SARS-CoV-2 by TMPRSS2-expressing cells. Proc Natl Acad Sci U S A. 2020;117(13):7001-3.
